# Proteome-wide Mendelian randomization analyses to identify potential therapeutic targets for lung function and respiratory disease

**DOI:** 10.1101/2025.02.07.25321860

**Authors:** Jing Chen, Nick Shrine, Kayesha Coley, Richard J. Packer, Ahmed Edris, Abril G. Izquierdo, Brandon Lim, Frank Dudbridge, Robin G Walters, Ian P Hall, Louise V Wain, Martin D Tobin, Anna L. Guyatt, SpiroMeta Consortium, CHARGE consortium

## Abstract

**RATIONALE:** Impaired lung function predicts mortality and is a diagnostic criterion for chronic obstructive pulmonary disease (COPD). Proteins are often the target of pharmacological interventions, therefore identifying causal links between proteins and lung function could inform understanding of COPD pathophysiology and suggest therapeutic targets. We aim to infer the potential impact of circulating protein levels on lung function, using strictly defined cis protein quantitative trait loci (cis-pQTLs) as genetic instrumental variables for Mendelian randomisation (MR).

**METHODS:** We applied two-sample MR by integrating protein GWAS data (2,923 proteins, 48,195 UK Biobank European participants) with lung function GWAS data (four lung function traits, 149,166 European participants from 36 non-UK Biobank cohorts). We selected strictly defined cis-pQTLs, within 100 kilobase pairs of a transcription start site and strongly associated (P≤5×10^−9^) with protein levels, and applied single-cis-MR analysis (Wald ratio method). Sensitivity analyses included colocalization analysis (to distinguish causal effects from genomic confounding by linkage disequilibrium), and bidirectional MR to explore possible reverse causation. Replication analysis was conducted where possible. We used the Drug-Gene Interaction Database and phenome-wide association studies (PheWAS) to inform biological and clinical interpretation of identified proteins.

**RESULTS:** We curated 1,841 proteins with a suitable cis-pQTL instrument, and evaluated evidence for causal effects of these proteins on four lung function traits. The single-cis MR analysis implicated 16 proteins for lung function at a Bonferroni-corrected threshold (Wald ratio estimator P<1.71×10*-5*), with evidence from colocalization. Of these, 10 proteins have been previously implicated either by lung function GWAS, or from other MR analyses with colocalization. Surfactant protein D (SFTPD) has been highlighted in previous respiratory MR analyses and variants in SFTPD have been previously reported to be associated with emphysema; our PheWAS suggested that this variant has a relatively specific effect on lung function as it was associated with no non-respiratory traits at a FDR<1%. In contrast to previous expression QTL evidence, our study suggested that ITGAV inhibition could reduce FEV*1*/FVC; we note that reduced lung function was also seen in a recent trial of an ITGAV inhibitor (NCT01371305). Our MR analysis implicated six proteins not implicated by previous lung function GWAS or MR (DTD1, PILRA, PTPRK, TDPRK, GRHPR, NUDT5).

**CONCLUSIONS:** Our protein-based approach identified proteins that may be causally related for lung function variability. We highlight known protein drug targets, and identify several new proteins which are potentially therapeutic targets but warrant further follow up for potential utility and safety.

## Introduction

Impaired lung function predicts mortality and is a diagnostic criterion for chronic obstructive pulmonary disease (COPD), the sixth leading cause of global disability-adjusted life years.^1^ Whilst beta-2 adrenoreceptor agonists, antimuscarinic agents, and corticosteroids are used in COPD, there has been a resurgence in drug development for COPD, with multiple clinical trials in progress for biologics and the first FDA-approved biologic, dupilumab.^2^ One approach to identifying drug targets involves the use of genetic association studies, which reported 1020 independent associations with lung function and COPD.^3–5^ Protein associations with lung function may identify drug targets as proteins are pharmaceuticals that often target proteins, and a proteomic-based score predicted COPD onset in UK Biobank (UKB)^6^. However, confounding and reverse causation typically affect such observational studies, which can result in non-causal protein-lung function associations being prioritised, and causal lung function associations being overlooked.

Integration of genomic and proteomic data enables inference of circulating protein associations with lung function.^7,8^ Genetic variants that act on levels of a specific protein (protein quantitative trait loci, pQTLs) are often located near the gene encoding that protein.^9^ These cis-pQTLs are often strongly associated with a protein level, with a direct and clear mechanism, making them suitable as instruments for Mendelian randomization to inform drug target identification.^7^

We aimed to integrate data on genomic variation and proteins in 50,395 UKB participants with data on genomic variation and lung function in 149,166 participants from 36 cohorts to estimate the predicted effect of protein levels on four lung function traits, via strictly defined cis-pQTLs. Whilst the proteomics of lung function has been studied in smaller sample sizes^10^ and limited numbers of traits,^11^ our study draws upon the largest GWAS of lung function. We discuss what is known about how the identified proteins may influence the pathophysiology of obstructive lung disease, and their potential utility (including known cautions) when considering these proteins as drug targets.

## Methods

### Genetic association analyses for proteomics

The UK Biobank Pharma Proteomics Project (UKB-PPP) characterised the plasma proteomic profiles of 54,306 UKB participants. Sample processing and proteomic measurement quality control in UKB-PPP is detailed elsewhere.^12^ We undertook genome-wide association analyses of 2,923 proteins in 50,395 participants, including 48,195 European (EUR) participants (EUR single cis-pQTL MR analyses), plus 1,202 African- (AFR), 482 American- (AMR), 237 East Asian- (EAS), and 729 South Asian- (SAS) ancestry participants in UK Biobank (ancestry described in^5^). Relative protein abundancies were log_2_- transformed, then adjusted using linear regression for age, sex and protein assay batch. GWAS were performed on untransformed protein residuals using REGENIE v3.2.6 via a two-step procedure to account for population structure,^13^ including covariates of UKB genetic array and the first 10 genetic principal components. For MR analyses, effect sizes of SNPs on proteins were converted to changes in standard deviations (SDs).

### Single-cis-MR analysis

Study design is explained in **Figure 1**. We selected strictly defined cis-pQTLs as the instruments for the MR analysis, defined as SNPs strongly associated (P≤5×10^−9^) with levels of a protein, and within 100 kilobases of the transcription start site (TSS) of a protein’s cognate gene. We applied two-sample MR by integrating the protein (exposure) EUR genetic association data from UKB with lung function trait (outcome) GWAS data (FEV_1_, FVC, FEV_1_/FVC and PEF) from meta-analyses of 36 non-UKB cohorts of 149,166 European participants. We implemented single-cis-MR analysis which utilized the strongest cis-pQTL variant (i.e. the SNP with lowest P value, where multiple variants met the criteria above) for each protein as the instrumental variable, and the Wald ratio^14^ to estimate the causal effect of each protein on lung function. Protein-lung function associations exceeding P<1.71×10^−5^ (Bonferroni correction for 2,923 proteins, of which 1,841 had cis-pQTLs) were prioritised for follow-up analyses. We do not report proteins encoded by genes in the human major histocompatibility complex (MHC) region (chromosome 6, 26Mb-34Mb) due to extensive linkage disequilibrium in this region. We report all coordinates using GRCh37/hg19.

**Figure 1.**
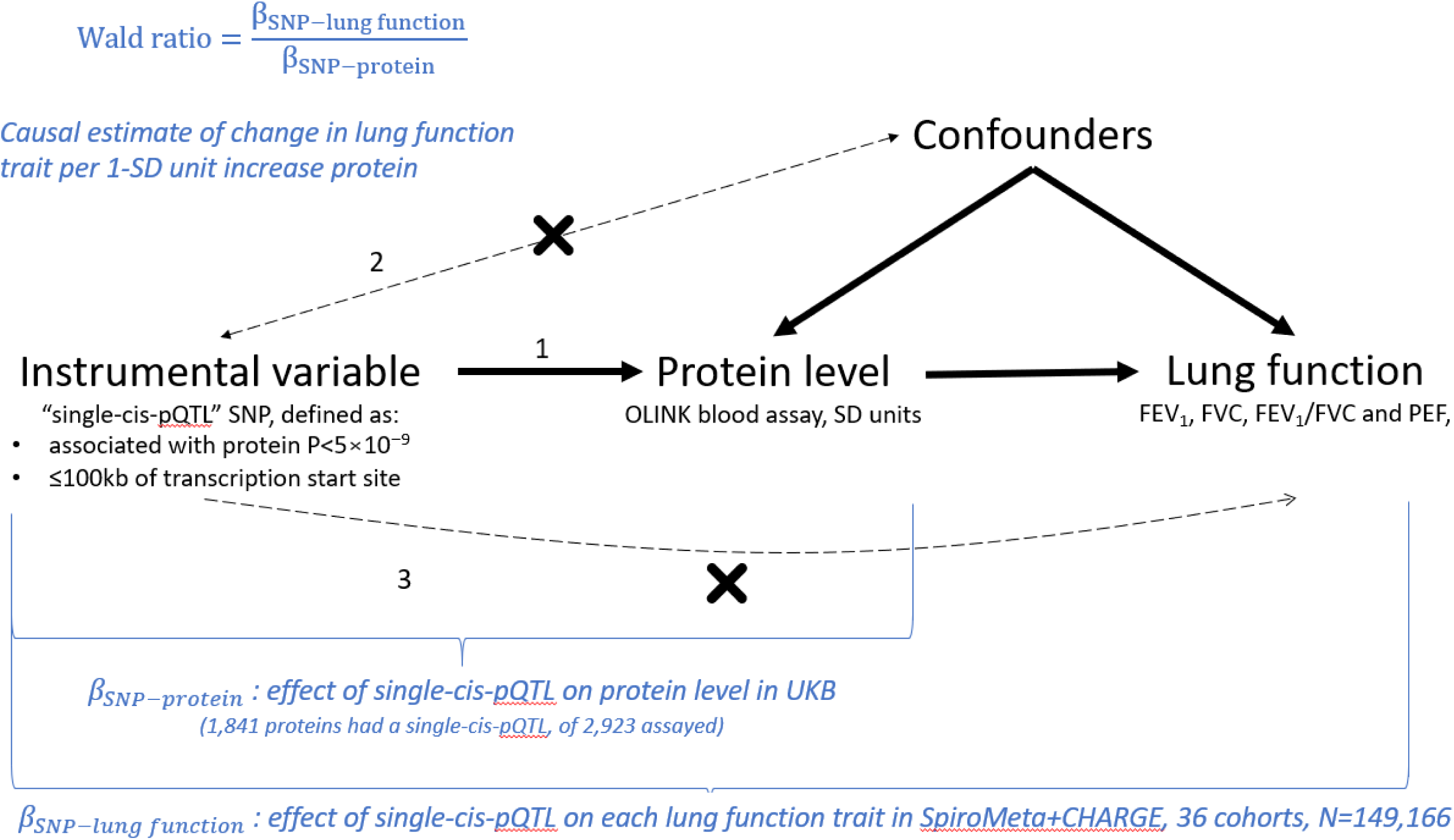
We performed two-sample MR analyses to estimate causal effects of 1,841 proteins on four lung function traits: forced expiratory volume in 1 second (FEV_1_), forced vital capacity (FVC), their ratio (FEV_1_/FVC) and the peak expiratory flow (PEF). The assumptions of MR are shown in the directed acyclic graph. MR is a form of instrumental variable analysis able to identify causal associations if: (1) the instrumental variable are associated with the exposure of interest (here, protein levels); (2) the instrumental variable is not associated with unobserved confounders of the exposure-outcome association (straight dashed arrow), and (3) the only association between the genetic variants and the outcome is via the exposure, and not via an alternate pathway (i.e., no ‘horizontal pleiotropy’, curved dashed arrow). As it is difficult to verify this last assumption, we focused this study on cis-pQTLs which are more likely to be valid instruments (PMID https://pubmed.ncbi.nlm.nih.gov/27342221/), since these variants are less likely to violate this assumption than trans-pQTLs. Strictly defined single-cis-pQTL SNPs were sought for use as instrumental variables (IVs) for 2,923 proteins (1,841 proteins had a valid IV). Causal estimates, as change in the lung function trait per 1-SD protein increase were calculated for each protein-lung function trait pair using a Wald ratio estimator, by dividing the effect on the IV on lung function (estimated in the SpiroMeta+CHARGE consortia, N=149,166) by the effect of the IV on protein level (estimated in UK Biobank EUR participants, N=48,195).

### Colocalization analysis

The prioritized MR results were evaluated by Bayesian colocalization.^15^ For genomic regions with multiple signals, we performed conditional analyses using GCTA-COJO^16^ using an LD reference panel of 10,000 randomly selected UK Biobank European ancestry participants. We prioritised MR findings for which the posterior probability (PP) of a shared signal was >0.7^17^ and where visual inspection of the mirror (“Miami”) plot was consistent with colocalization of the pQTL and lung function GWAS signal (**Supplementary Figure 1** and **Supplementary Table 1**).

### Reverse causation test of single-cis-MR findings

To explore possible reverse causation, i.e. whether a protein level could be causally influenced by the lung function trait under study, we performed MR in the opposite direction.^18^ For each protein identified in the protein➔lung function MR analysis, we chose the lung function trait for which there was strongest evidence of causality by that protein. We identified independent lung function signals for the relevant trait from Shrine et al.^5^ (independent SNPs associated at P<5×10^−9^), and applied Steiger filtering,^19^ removing SNPs which explained more variance in the protein levels than the lung function trait. We used the remaining SNPs as instruments for the lung function trait, and used the inverse variance-weighted (IVW) MR method (as implemented by mr_ivw() in the Two-Sample MR package) to estimate the causal effect of that lung function trait on the relevant protein.

### Respiratory-wide association study

To determine the clinical relevance of the causal associations identified with lung function, we performed a respiratory phenome-wide association study (PheWAS), querying each cis-pQTL against 28 GWASs (**Supplementary Table 2**) for: asthma, bronchiectasis, bronchopneumonia, chronic bronchitis, COPD, chronic sputum production, emphysema, idiopathic pulmonary fibrosis (IPF), respiratory infections, interstitial lung abnormalities (ILA) and pneumothorax. We report associations with p-value <0.01.

### PheWAS

For proteins identified in the single-cis MR analysis, we undertook single-variant PheWAS for the cis-pQTLs selected as instruments across 1,909 traits in up to 430,402 unrelated EUR individuals in the UK Biobank using Deep-PheWAS (PMID 36744935). The PheWAS results were aligned to protein-increasing alleles. We report associations with FDR<1%.

### Replication analysis

Proteins identified from the single-cis-MR analysis were further investigated in a replication analysis using the Somalogic-derived pQTL data from deCODE (PMID: 34857953) and INTERVAL (PMID: 29875488) studies. We applied the same two-sample MR approach by integrating pQTL data for proteins available from either deCODE or INTERVAL with the lung function trait GWAS data from UK Biobank European population. Protein-lung function associations that met a Bonferroni-corrected significance threshold for the number of proteins prioritised from the MR were considered replicated.

### Druggability

We queried the gene-drug interaction table from the Drug-Gene Interaction Database for each prioritised protein; associated drugs were linked to ChEMBL IDs, and indications annotated using MeSH headings.

### Associations of top proteins in diverse ancestries

For proteins prioritised by the single cis-pQTL MR analyses in EUR participants, we explored whether the same cis-pQTL was associated at P<0.05, within ancestry subgroups defined in.^5^

## Results

Our GWAS studies of 2,923 proteins identified 1,841 proteins with a suitable cis-pQTL instrument (SNP-protein P≤5×10^−9^ and <100kb of TSS). The cis-pQTL MR analyses causally implicated 16 proteins in lung function (p<1.71×10^−5^, Bonferroni correction for 2,923 proteins), including seven proteins not implicated by previous GWAS – i.e. where the pQTL SNP was independent of reported lung function signals (CCND2, DTD1, PILRA/PILRB, PTPRK, TDRKH, GRHPR, NUDT5) (**Figure 2**, **Supplementary Tables 3 and 4**).^5^. Nine of the proteins identified were causally implicated in lung function by SNPs that were in linkage disequilibrium with known lung function signals (COL6A3, EFEMP1, ITGAV, PRSS53, SFTPD, SLMAP, TNFRSF6B, TOP2B, WASHC3)^20^. Of the 16, eight proteins (CCND2, COL6A3, EFEMP1, ITGAV, SFTPD, SLMAP, TNFRSF6B and WASHC3) have also been reported as having evidence from Mendelian randomization, plus colocalization.^10,11^

**Figure 2.**
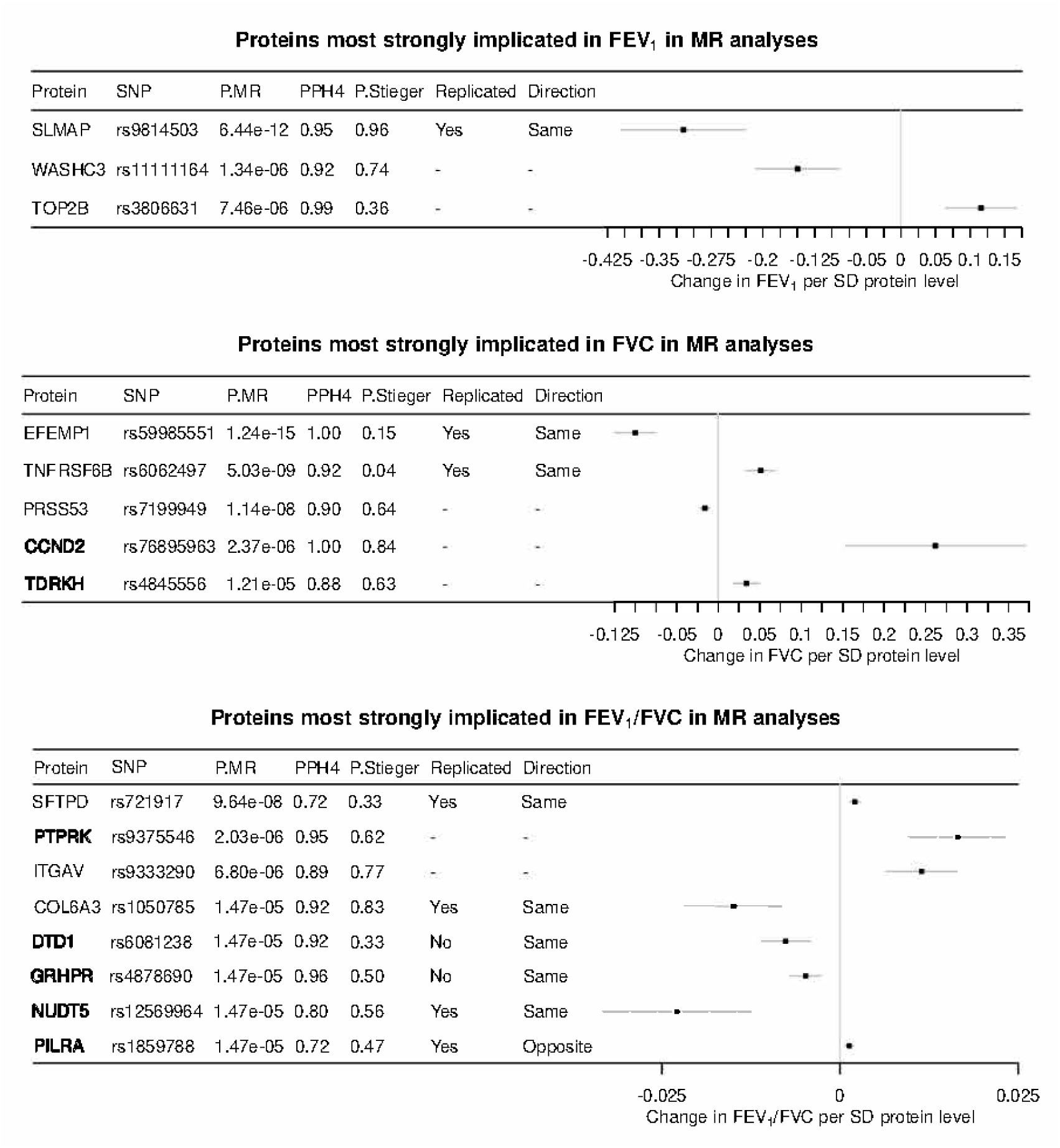
The 16 proteins implicated by the single-cis-MR analyses for the lung function traits. Forest plots of the Wald ratio estimates for the strongest associated lung function trait are shown, along with the p value for the Wald ratio method (P.MR), the posterior probabilities of colocalization (PPH4), the p value for the reverse causation test (P.Steiger) and replication results (Replicated and Direction, with a “-“ indicating that replication data were not available). Proteins which have not been previously implicated by a lung function signal are highlighted in bold. Boxes of the forest plot represent effect sizes, whiskers are 95% CIs. FEV_1_, forced expiratory volume in 1s; FVC, forced vital capacity; MR, Mendelian randomisation; SNP, single-nucleotide polymorphisms; SD, standard deviation.

For 12/16 proteins (CCND2, DTD1, EFEMP1, ITGAV, PILRA/B, PRSS53, PTPRK, SLMAP, TDKRH, TNFRSF6B, TOP2B, WASHC3), the cis-pQTL showed association (P<0.01) with at least one of the following phenotypes in the respiratory PheWAS: COPD, emphysema, asthma, IPF onset or diffusing capacity for carbon monoxide (DLCO), ILA, or chronic sputum production (see **Supplementary Table 5** for association results, and **Supplementary Table 6** for SNP-trait pairs missing in the respiratory PheWAS). Although no associations with SFTPD were observed in the respiratory PheWAS, its pQTL was previously reported as associated with emphysema.^20^

For FEV_1_ the strongest association was seen for the sarcolemma-associated protein, SLMAP. An increase in SLMAP predicted by rs9814503 was associated with reduced FEV_1_ (P= 6.44×10^−12^), with weaker associations with reduced FEV_1_/FVC (P=6.33×10^−5^) and reduced FVC (P=1.05×10^−5^). The SLMAP-increasing allele (C) of rs9814503 was associated with increased risk of asthma, childhood-onset asthma and COPD (**Supplementary Tables 2, 5 and 6**) in the respiratory PheWAS, with reduced diastolic blood pressure in the PheWAS (**Supplementary Table 7**), and SNPs in this gene (including rs17666667-T, r^2^=0.99 with rs9814503-C) are associated with reduced smoking initiation.^21^ We note that the C allele of rs9814503 is also associated with reduced FEV_1_, FVC, FEV_1_/FVC, and PEF in never smokers, therefore the association of this SNP with these lung function traits is not via its association with smoking.^5^ We previously showed a consistent FEV_1_ association in children (rs9872373-C [correlated with C allele of rs9814503, r^2^=0.98], −0.015, SE=0.004, P=0.0006).^5^ This pattern of association with phenotypes at different stages of life could be consistent with a mechanism influencing airflow obstruction in early life, then tobacco avoidance and fixed airflow obstruction in later life.

Additional predicted protein-FEV_1_ associations were seen for WASHC3 (WASH Complex Subunit 3) and TOP2B (DNA Topoisomerase II Beta). pQTLs showed association with decreased sputum production for WASHC3 and reduced risk of COPD for TOP2B.

For FVC, we observed the strongest association with EFEMP1, EGF Containing Fibulin Extracellular Matrix Protein 1. An increase in EFEMP1 predicted by rs59985551 was associated with lower FVC (P=1.24×10^−15^) (see also ^10^), increased FEV_1_/FVC (P=1.52×10^−7^), and decreased risk of emphysema. EFEMP1 is associated with elastic fibre formation and in the PheWAS, consistent with effects on reduced tissue elasticity, the EFEMP1-increasing allele rs59985551 (T) showed association with reduced height, reduced risk of hernias, reduced diverticular disease, and increased carpal tunnel syndrome.

TNFRSF6B is a member of the tumour necrosis factor receptor superfamily, and acts as a decoy receptor that can neutralise cytotoxic ligands. An increase in TNFRSF6B predicted by rs6062497 was also associated with increased FVC (P=5.03×10^−9^) and FEV_1_ (P=2.26×10^−8^) (see also ^10^), and the protein-increasing allele (C) was associated with a multiple respiratory traits, including reduced risk of asthma, COPD, IPF and bronchitis, and increased risk of pneumothorax. Notably, rs6062497 is downstream of RTEL1 (telomere elongation helicase), and telomere genes are known to play a role in the risk of interstitial lung diseases, for which FVC is the trait of primary relevance.^22^ In PheWAS, the protein-increasing allele is also associated with lower granulocytes (eosinophils and neutrophils), as well as lower CRP, reduced smoking heaviness and risk of smoking onset, reduced risk of asthma, and reduced risk of IBD. The same allele was associated with an increased risk of dental disorders and traits related to tissue laxity, such as diverticulosis.

For PRSS53 (serine protease 53), an increase in levels was instrumented by rs7199949, and associated with similar decreases in FVC (P= 1.14×10^−8^) and FEV_1_ (P=1.46×10^−8^). The PRSS53-increasing allele was also associated with increased risk of asthma, especially in children, and COPD. In PheWAS, the same allele was associated with many traits, including but not limited to those relating to increased body size, altered red cell traits (which might explain an association with increased HbA1C in the absence of association with diabetes-related traits), reduced diverticular disease, protein levels (including albumin), reduced vitamin D, and lipids.

An increase in CCND2 (G1/S-specific cyclin-D2, fundamental to the cell cycle) predicted by rs76895963 was associated with higher FVC (P=2.37×10^−6^). In the respiratory PheWAS, the protein-increasing allele (G) was associated with increased risk of COPD and ILA and reduced DLCO in IPF. PheWAS showed that the protein-increasing allele was also associated with growth-related traits, including increased height and weight, plus reduced type 2 diabetes risk.

TDRKH (Tudor And KH Domain Containing) was associated with increased FVC (P=1.21×10^−5^) and more weakly with increased FEV_1_ (P=0.00032), and the protein-increasing allele (A) was associated with reduced childhood-onset asthma (P=3.4 x 10^−9^) in the respiratory PheWAS and with reduced gamma-glutamyltransferase in the broader PheWAS.

The protein-increasing allele (A) of a non-synonymous (Met11Thr) SNP in SFTPD (rs721917, surfactant protein D) was associated with increased FEV_1_/FVC. SFTPD has been previously implicated in MR analyses of COPD,^10,23^ and variants in SFTPD have been reported to be associated with emphysema^20^ – our analyses corroborate these results in a disease-relevant quantitative trait measured in a general population, and we do not find evidence that this is driven by reverse causation. Our PheWAS results suggest that this variant has a relatively specific effect on lung function, with no non-respiratory traits associated in the PheWAS at an FDR<1%. We also found modest associations (0.01<P<0.05) of the protein-increasing allele (A) with decreased COPD, chronic bronchitis, emphysema, respiratory infections and asthma that support previous findings^23^ and the same allele is reported to be protective against severe COVID-19.^24,25^ SFTPD is involved in promotion of innate immunity, as well as surfactant regulation and lipid homeostasis.^23^ We observed consistent directions of effect for the SNP on SFTPD levels in UK Biobank (OLINK assay), and in China Kadoorie Biobank (OLINK and Somascan assays), but the same protein-increasing allele decreased FEV_1_/FVC in deCODE.^26^

PTPRK (protein tyrosine phosphatase receptor type K) regulates cell contact and adhesion, and negatively regulates the EGFR signalling pathway. The protein-increasing allele was associated with improved FEV_1_/FVC and reduced asthma risk, but with increased risk of abdominal aortic aneurysm, suggesting caution in considering PTPRK as a druggable target.

Our MR results for ITGAV (Integrin Subunit Alpha V) provide causal evidence in the opposite direction to previous eQTL evidence. ITGAV is a known druggable target (See **Supplementary Table 8**), and in the current analysis, the protein-increasing allele (rs9333290-G) was associated with increased FEV_1_/FVC and PEF, lower asthma, COPD and emphysema risk, reduced height, and increased phosphate. In our 2019 and 2023 GWAS of lung function, the allele associated with increased ITGAV gene expression^27^ was associated with lower FEV_1_/FVC (and higher FVC), at that time suggesting that α vβ 6 integrin inhibitors might have a beneficial effect in COPD. A trial of an α vβ 6 integrin inhibitor (NCT01371305) reported increased pulmonary fibrosis exacerbations in the treatment group and lung function decline at higher doses;^28^ the drug has not progressed further. These clinical trial findings are consistent with our pQTL findings (and work presented in ^10^), and our work highlights that eQTL and pQTL results do not always tally.^29^

The Collagen Type VI Alpha 3 Chain (COL6A3) is found in most connective tissues. We found evidence that higher COL6A3 levels, as predicted by rs1050785-A, were causally associated with lower FEV_1_/FVC (P=1.47×10^−5^), but the same SNP was not associated with any clinical phenotype at p<0.01 in the respiratory PheWAS. The only drugs identified in our druggability analyses targeting COL6A3 included a collagenase and recombinant protease, ocriplasmin, neither of which we would hypothesise as being tractable drug targets for respiratory disease.

DTD1 (D-Aminoacyl-TRNA Deacylase 1) is involved in initiating DNA replication. The allele (G) associated with increased DTD1 was associated with lower FEV_1_/FVC, increased COPD risk in the respiratory PheWAS and with raised monocytes and creatinine levels in the broader PheWAS. Higher GRHPR (Glyoxylate And Hydroxypyruvate Reductase) and NUDT5 (Nudix Hydrolase 5) levels were both associated with lower FEV_1_/FVC; however, neither of the SNPs that instrumented these proteins were associated (P<0.01) with any clinical respiratory trait. The NUDT5 cis-pQTL, rs12569964, was associated with height and type-2 diabetes traits, but no PheWAS associations were observed for GRHPR at FDR<1%.

The cis-pQTL for paired immunoglobin like type 2 receptor alpha (PILRA) and beta (paralog PILRB), involved in immune system signal transduction and herpes simplex virus cellular entry, was a non-synonymous SNP, rs1859788. Our MR analyses showed an association between increased PILRA and increased FEV_1_/FVC and similarly, an association between PILRB and increased FEV_1_/FVC. The PILRA/PILRB-increasing allele (G) was associated with lower risk of emphysema, COPD and asthma and notably increased chronic sputum production in the respiratory PheWAS. However, in the broader PheWAS, the protein-increasing allele (G) was associated with an increase in HbA1c; this allele has been also reported as associated with an increased risk of dementia ^30^, which could caution against PILRA or PILRB as respiratory drug targets.

Of the 16 proteins we highlight, 9 were available to assess replication in Somalogic-derived proteomics resources (COL6A3, DTD1, EFEMP1, GRHPR, NUDT5, PILRA, SFTPD, SLMAP, TNFRSF6B) (**Supplementary Table 9**). For 7 of these 9 proteins (SLMAP, PILRA, COL6A3, EFEMP1, NUDT5, SFTPD, TNFRSF6B), MR-derived protein-lung function associations met our replication threshold (P<3.13×10^−3^, correcting for 16 proteins, **Supplementary Table 9**). One of these proteins, NUDT5, replicated and has not been implicated by a previous lung function signal, and although noted in^10^, that publication did not find evidence of colocalization (and thus was not reported as a main result). For PILRA (predicted by a non-synonymous SNP), the MR-derived protein-lung function association in the Somalogic-derived replication analyses had an opposite direction of effect (the protein-increasing allele reduced FEV_1_/FVC). For the remaining two proteins (DTD1, GRHPR) that did not reach significance in the replication analysis, we observed a consistent effect direction.

For the 16 proteins we highlight (counting PILRA/PILRB as one protein), we explored whether the top cis-pQTL for each protein was strongly associated in four other ancestry subgroups in UK Biobank (AFR, AMR, EAS, and SAS). For the AFR, AMR, EAS and SAS subgroups respectively, the number of SNPs (out of the SNPs available for that subgroup, maximum N=16) significant for their respective protein level at P<0.05 was 8/16 for AFR (N=1,185), 9/15 for AMR (N=479), 2/9 for EAS (N=235), and 8/15 for SAS (N=719) (see **Supplementary Table 10**). Of note, the cis-pQTL SNPs for PILRA/PILRB and SFTPD were available in all five subgroups, and were strongly associated with these protein levels in all subgroups. Importantly, we note that the SNPs for these proteins were missense SNPs, meaning that they may be more likely to be the causal variant driving protein levels.

## Discussion

We integrated genomic and proteomic data from UK Biobank to study the causal role of proteins on lung function traits measured by spirometry. Instead of using observational epidemiological associations to prioritise protein-outcome pairs for study, we applied a hypothesis-free agnostic approach, identifying cis-pQTLs for as many proteins as possible that had been measured across the genome, using a strict definition (p<5×10^−9^, SNP <100kb from a transcription start site). We identified 16 proteins with evidence of causality on lung function, of which seven had not been previously identified in large-scale lung function GWAS. Eight of the 16 proteins (CCND2, COL6A3, EFEMP1, ITGAV, SFTPD, SLMAP, TNFRSF6B and WASHC3) have also been identified in other recent MR efforts with colocalization.^10,11^ Our approach confirms the utility of a ‘genetics-first’ approach for identifying protein-disease associations, and provides a list of proteins amongst which some may be worthy of exploration as therapeutic targets.

Our results also highlight the qualitative nature of evidence that cis-pQTL evidence can add. For example, the evidence for the effect on ITGAV on lung function is conflicting when results of eQTL and pQTL data are compared. In a previous paper, we found that increased ITGAV expression was associated with decreased lung function,^4^ yet subsequent trials have reported that inhibition of this molecule led to pulmonary fibrosis exacerbations and lung function decline.^28^ Our current MR results (see also ^10^) would have predicted the results of this trial, and yet we have previously hypothesised the opposite.^4^

We have previously shown that studying lung function is a well-powered way of studying relevant chronic respiratory diseases, such as COPD and IPF.^5,31^ To explore the relevance of protein-lung function associations to clinical disease phenotypes, we performed a ‘respiratory-wide association study’ (Respiratory PheWAS), and found that many of our cis-pQTL SNPs were associated with a clinically-relevant outcome, such as asthma, COPD (and sub-components emphysema and bronchitis), interstitial lung disease, and clinically-relevant symptoms such as sputum production.

Whilst our results suggest a causal role for all of the proteins we prioritise, their suitability as therapeutic targets varies. Some proteins, such as WASHC3 and TOP2B, have such fundamental roles that they seem implausible as therapeutic targets to improve lung function – indeed topoisomerase inhibitors such as etoposide exist, but as chemotherapeutic agents, thus with low therapeutic indices and wide-ranging effects that would render them unsuitable for repurposing. Other proteins we and others^10,23^ implicate, such as SFTPD, have a known role in the lung, are overexpressed in pulmonary tissue and had few off-target predicted effects in our PheWAS, adding to the evidence base of SFTPD as a possible therapeutic target. ITGAV had opposite directions of predicted effects from eQTL and pQTL, highlighting that triangulating evidence from multiple sources is important predict effects of target modulation. Finally, our PheWAS approaches provide corroborative insight into the biology of some of the putative targets suggested. For example, the PheWAS suggests that inhibiting EFEMP1^10^ could improve FVC and reduce risk of pulmonary fibrosis, but increase risk of traits related to tissue laxity. Thus, integrating PheWAS data is helpful for understanding potential off-target effects, whether deleterious or not.

Strengths of our work include our strict cis-pQTL definition to mitigate against risk of horizontal pleiotropy (that is, effects of the SNP on the outcome that do not pass via the protein of interest, which bias MR effect estimates, see Figure 1), and requirement for corroborating evidence from colocalization analysis.^32^ By definition, the cis-pQTLs not implicating a known lung function signal must not be genome-wide significant for lung function. However, we would argue that these cis-pQTLs could have a higher prior probability of association with lung function than a random SNP. Instruments for a protein that are i) strong and ii) have a highly plausible and specific role on the protein of interest are useful in MR.^7^ However, at the same time, the strictness of definition will have limited our ability to identify IVs for as many proteins, had we relaxed this criterion. Since we hypothesised that any protein could be causal for lung function, our discovery Bonferroni correction threshold incorporated the number of proteins assayed, i.e. stricter than only considering the number of proteins for which we found cis-pQTLs. Despite this, we acknowledge that the pool of proteins measured by the UKP-PPP project is only a subset of the human proteome. Others have advocated a stricter discovery alpha threshold, i.e. P<0.005, for hypothesis-free analyses, yet we would argue that the proteins assayed by UKB-PPP are not a random sample of the proteome, and thus may have a higher prior probability of causality. We used a very large resource of proteomics data, measured using an assay robust to aptamer effects, and the largest set of lung function GWAS results available (without sample overlap – which can bias MR results in an anti-conservative manner). We also assessed for evidence of reverse causation. However, we used discovery SNP-protein results from a European only subset, as sample sizes for other ancestry groups were small. We observed concordant associations across all ancestry subgroups for some proteins proxied by missense SNPs, variants which may be more likely to be the causal driver of protein levels, yet may also be due to changes on protein structure. Yet, absence of concordant associations for other SNP-protein pairs across other ancestries may reflect some combination of SNP availability, statistical power, and whether or not the cis-pQTL IV is in linkage disequilibrium with the causal SNP in non-EUR populations. Another strength is that we were able to explore corroborative evidence for over half of our prioritised proteins by using independent replication from other sources. As proteomics resources grow, we may find replication for the remaining proteins. However, since Olink protein assays are antibody-based and Somalogic assays are aptamer-based, for non-synonymous SNPs, these could influence antibody and aptamer binding in different ways, making interpretation more challenging.

To conclude, we used a genome-wide, proteome-wide approach, agnostic to observational epidemiological associations, to identify proteins that may be potentially causal for lung function. We were able to replicate the vast majority of these, and highlight a role for the SNPs instrumenting them in risk of relevant clinical phenotypes. Together with other sources of information to inform efficacy and safety, our findings could inform the choice of therapeutic targets for chronic lung diseases such as COPD.

## Data Availability

All data produced in the present study are available upon reasonable request to the authors

## Supplementary Figure and Table Legends

**Supplementary Table 1**

Bayesian colocalization between SNP-protein and SNP-lung function trait associations. Colocalisation was undertaken for all non-HLA cis-pQTLs predicting proteins that were found to have evidence of causality in single cis-pQTL MR analyses. Summary statistics for common variants (minor allele frequency > 0.01) located within 1 Mb of the cis-pQTL from lung function GWAS and pQTL data were extracted for statistical inference. Colocalisation was undertaken for multiple traits where multiple traits passed Bonferroni correction for the MR analyses. Protein = protein predicted by cis-pQTL. SNP = chromosome, position (GRCh37), alleles (see **Supplementary Table 3** for details of protein-increasing allele); PPH4.coloc.abf = posterior probability of hypothesis 4, e.g. the SNP-protein and SNP-lung function trait signal are the same, using the approximate Bayes Factor colocalization analyses under single causal variant assumption (standard coloc method); PPH4.conditional = posterior probability of hypothesis 4, using conditional analysis and approximate Bayes Factor colocalization analyses, for regions failed by coloc.abf which may have multiple signals within the region (conditional coloc method).

**Supplementary Table 2**

Sources of data used in the respiratory-wide association study (RespWAS).

**Supplementary Table 3**

Results of Wald ratio Mendelian randomization (MR) for 21 proteins (including two that did not show evidence of colocalization). Results are shown for the lead lung function trait (i.e. with the lowest P-value in MR analyses of that protein). StrongestTrait = most significant lung function trait (out of FEV1, FVC, FEV1/FVC and PEF) for each MR analysis of a given protein. Consequence = cis-pQTL SNP annotation (using ANNOVAR^33^); PPH4 (coloc) = highest posterior probability (standard or conditional) of colocalization hypothesis 4; SE = standard error; SD = standard deviation; LOG10P = −log_10_ P-value; LOG10P.Steiger = −log_10_ P-value for Steiger test; Effect alleles are the protein-increasing alleles; Novel LF signal = cis-pQTL not in linkage disequilibrium (r^2^ < 0.2) with known lung function signal. Non-novel signals defined as cis-pQTL in linkage disequilibrium (r^2^ > 0.2) with a known lung function signal. LD (r^2^) = the linkage disequilibrium measurement r^2^ between cis-pQTL and the previously reported lung function signal.

**Supplementary Table 4**

Results of Wald ratio Mendelian randomization (MR) for 21 proteins (including two that did not show evidence of colocalization). Results are shown for all four lung function traits. StrongestTrait = most significant lung function trait (out of FEV1, FVC, FEV1/FVC and PEF) for each MR analysis of a given protein; Consequence = cis-pQTL SNP annotation (using ANNOVAR^33^); PPH4 (coloc) = highest posterior probability (standard or conditional) for colocalization hypothesis 4; SE = standard error; SD = standard deviation; LOG10P = −log_10_ P-value; Steiger = estimates for Steiger test (see Methods); Effect alleles are the protein-increasing alleles. The column ‘Associated with coded gene’ denotes whether the cis-pQTL is annotated as a coding variant for the protein reported.

**Supplementary Table 5**

Results of respiratory-wide association study (RespWAS). Each set of GWAS results interrogated has a unique ID (RespPheWAS_ID). Details of the study are given in columns explaining the phenotype and subcategories, the cohort analysed and the ancestry. Later columns then detail the SNP extracted from this study, including its unique rsID, chromosomal location, alleles (included the coded ‘effect_allele’, to which the effect estimate refers), sample size, and summary statistics for the association of this SNP with the trait in question.

**Supplementary Table 6**

This table details SNP-trait associations which were missing from the RespWAS, including details of the GWAS interrogated (trait studied, cohort name, ancestry, and sample size).

**Supplementary Table 7**

Results for phenome-wide association study (PheWAS), conducted using DeepPheWAS software.^34^ Results are shown for single-variant PheWAS results for each cis-pQTL. EUR = European; collective_name = nearest gene name; category = type of outcome; N_ID = sample size; FDR = false discovery rate; P = P value; OR = odds ratio; L95 = lower boundary of the 95% confidence interval; U95 = upper boundary of the 95% confidence interval; MAF = minor allele frequency; MAC = minor allele count; MAC_cases = minor allele count in cases; MAC_controls = minor allele count in controls; Z_T_STAT = the Z or T STAT output from Plink2; SE = standard error.

**Supplementary Table 8**

The prioritised proteins were interrogated against the gene-drug interaction table from the Drug-Gene Interaction Database (DGIDB). Associated drugs were linked to ChEMBL IDs, and indications annotated using MeSH headings. Protein = prioritised protein; Drug = drug/compound name; Indication(Phase); drug indication and clinical trial phase (1-4); ChEMBL_ID; drug/compound identification number from ChEMBL; MAB = whether the drug is a monoclonal antibody; Cancer; whether the drug is used to for the treatment of some form of cancer.

**Supplementary Table 9**

Results of replication analysis for the proteins identified from the single-cis-MR analysis. Single-cis-MR (Wald ratio MR) results are shown for 9/16 proteins available for analysis in all four lung function traits. Source indicate the data source used for pQTL replication; SE = standard error; P_min = the minimum P value of Wald ratio MR from all the lung function traits.

**Supplementary Table 10**

Results of cis-pQTLs association with proteins in different ancestry groups in UK Biobank. AFR = African; AMR = American/Hispanic; EAS = East Asian; EUR = European; SAS = South Asian; ALLELE0 = non-effect allele; ALLELE1 = effect allele; A1FREQ = allele frequency for the effect allele; INFO = imputation quality score for the genetic variant; N = sample size; BETA = effect size; SE = standard error; LOG10P = −log_10_ P-value; P = P-value.

**Supplementary Figure 1**

Mirror plots to compare the region plots of protein-genetic associations with those of lung function-genetic associations for visualization of colocalization. Each point is a genetic variant. Points above the x axis show the strength of association with the lung function trait and points below the x axis show the strength of association with the protein level. Variants are coloured by their linkage disequilibrium with the cis-pQTL.

## Acknowledgements

The research was conducted using UK Biobank, under applications 648 and 43027.

This research used the ALICE and SPECTRE High Performance Computing Facilities at the University of Leicester.

## References

1. Vos, T. et al. Global burden of 369 diseases and injuries in 204 countries and territories, 1990-2019: a systematic analysis for the Global Burden of Disease Study 2019. The Lancet 396, 1204–1222 (2020).

2. Mullard, A. FDA approves first monoclonal antibody for COPD. Nat Rev Drug Discov 23, 805 (2024).

3. Sakornsakolpat, P. et al. Genetic landscape of chronic obstructive pulmonary disease identifies heterogeneous cell-type and phenotype associations. Nat Genet 51, 494–505 (2019).

4. Shrine, N. et al. New genetic signals for lung function highlight pathways and chronic obstructive pulmonary disease associations across multiple ancestries. Nat Genet 51, 481–493 (2019).

5. Shrine, N. et al. Multi-ancestry genome-wide association analyses improve resolution of genes and pathways influencing lung function and chronic obstructive pulmonary disease risk. Nat Genet 55, 410–422 (2023).

6. Gadd, D.A. et al. Blood protein levels predict leading incident diseases and mortality in UK Biobank. medRxiv, 2023.05.01.23288879 (2023).

7. Swerdlow, D.I. et al. Selecting instruments for Mendelian randomization in the wake of genome-wide association studies. International Journal of Epidemiology 45, 1600–1616 (2016).

8. Davey Smith, G. & Hemani, G. Mendelian randomization: genetic anchors for causal inference in epidemiological studies. Hum Mol Genet 23, R89–98 (2014).

9. Fauman, E.B. & Hyde, C. An optimal variant to gene distance window derived from an empirical definition of cis and trans protein QTLs. BMC Bioinformatics 23, 169 (2022).

10. Aggarwal, M. et al. Plasma Protein Biomarkers of Spirometry Measures of Impaired Lung Function. Chest (2024).

11. Su, C.-Y. et al. Multi-ancestry proteome-phenome-wide Mendelian randomization offers a comprehensive protein-disease atlas and potential therapeutic targets. medRxiv, 2024.10.17.24315553 (2024).

12. Sun, B.B. et al. Plasma proteomic associations with genetics and health in the UK Biobank. Nature 622, 329–338 (2023).

13. Mbatchou, J. et al. Computationally efficient whole-genome regression for quantitative and binary traits. Nat Genet 53, 1097–1103 (2021).

14. Lawlor, D.A., Harbord, R.M., Sterne, J.A., Timpson, N. & Davey Smith, G. Mendelian randomization: using genes as instruments for making causal inferences in epidemiology. Stat Med 27, 1133–63 (2008).

15. Giambartolomei, C. et al. Bayesian test for colocalisation between pairs of genetic association studies using summary statistics. PLoS Genet 10, e1004383 (2014).

16. Yang, J. et al. Conditional and joint multiple-SNP analysis of GWAS summary statistics identifies additional variants influencing complex traits. Nat Genet 44, 369–75, s1-3 (2012).

17. Zhang, L.Y. et al. Systematic Druggable Genome[Wide Mendelian Randomization Identifies Therapeutic Targets for Functional Outcome After Ischemic Stroke. Journal of the American Heart Association 13, e034749 (2024).

18. Timpson, N.J. et al. C-reactive protein levels and body mass index: elucidating direction of causation through reciprocal Mendelian randomization. Int J Obes (Lond) 35, 300–8 (2011).

19. Hemani, G., Tilling, K. & Davey Smith, G. Orienting the causal relationship between imprecisely measured traits using GWAS summary data. PLoS Genet 13, e1007081 (2017).

20. Ishii, T. et al. Involvement of surfactant protein D in emphysema revealed by genetic association study. Eur J Hum Genet 20, 230–5 (2012).

21. Saunders, G.R.B. et al. Genetic diversity fuels gene discovery for tobacco and alcohol use. Nature 612, 720–724 (2022).

22. Allen, R.J. et al. Genome-Wide Association Study of Susceptibility to Idiopathic Pulmonary Fibrosis. Am J Respir Crit Care Med 201, 564–574 (2020).

23. Obeidat, M. et al. Surfactant protein D is a causal risk factor for COPD: results of Mendelian randomisation. Eur Respir J 50(2017).

24. Salvioni, L. et al. Surfactant protein D (SP-D) as a biomarker of SARS-CoV-2 infection. Clin Chim Acta 537, 140–145 (2022).

25. A first update on mapping the human genetic architecture of COVID-19. Nature 608, E1–e10 (2022).

26. Ferkingstad, E. et al. Large-scale integration of the plasma proteome with genetics and disease. Nature Genetics 53, 1712–1721 (2021).

27. Jansen, R. et al. Conditional eQTL analysis reveals allelic heterogeneity of gene expression. Hum Mol Genet 26, 1444–1451 (2017).

28. Raghu, G. et al. Randomized Phase IIa Clinical Study of an Anti-α(v)β(6) Monoclonal Antibody in Idiopathic Pulmonary Fibrosis. Am J Respir Crit Care Med 206, 1166–1168 (2022).

29. Pietzner, M. et al. Mapping the proteo-genomic convergence of human diseases. Science 374, eabj1541 (2021).

30. Rathore, N. et al. Paired Immunoglobulin-like Type 2 Receptor Alpha G78R variant alters ligand binding and confers protection to Alzheimer’s disease. PLoS Genet 14, e1007427 (2018).

31. Allen, R.J. et al. Longitudinal lung function and gas transfer in individuals with idiopathic pulmonary fibrosis: a genome-wide association study. The Lancet Respiratory Medicine 11, 65–73 (2023).

32. Zheng, J. et al. Recent Developments in Mendelian Randomization Studies. Curr Epidemiol Rep 4, 330–345 (2017).

33. Wang, K., Li, M. & Hakonarson, H. ANNOVAR: functional annotation of genetic variants from high-throughput sequencing data. Nucleic Acids Res 38, e164 (2010).

34. Packer, R.J. et al. DeepPheWAS: an R package for phenotype generation and association analysis for phenome-wide association studies. Bioinformatics 39(2023).

